# Attachment-related anxiety and social anxiety: the mediating role of self-esteem

**DOI:** 10.1101/2024.05.28.24308030

**Authors:** Jacob Lincoln, Liesbeth Muriel Tip, Sofia De La Fuente Garcia

**Affiliations:** NHS Highland; Mindler UK; Department of Clinical Psychology, School of Health in Social Sciences, University of Edinburgh

## Abstract

Numerous studies have found an association between attachment-related anxiety and social anxiety. However, none have investigated the potential role of the internal working model of the self in explaining this relationship. The purposes of this study were to replicate the finding that attachment-related anxiety and social anxiety are associated, and to test whether the internal working model of the self mediated this relationship. The internal working model of the self was operationalised by measuring self-esteem. It was hypothesised that attachment-related anxiety, self-esteem, and social anxiety would be intercorrelated, and that self-esteem would mediate the relationship between attachment-related anxiety and social anxiety. A sample of 63 adults (79.4% female) was recruited through social media, University course groups, and snowball sampling. Participants completed an online survey that consisted of a reduced version of the anxiety subscale of the Experiences in Close Relationships-Revised Scale, the Rosenberg Self-Esteem Scale, and the Social Interaction Anxiety Scale. Ethical approval was given by the University of Edinburgh School of Health in Social Science Research Ethics Committee. Pearson correlation tests showed that attachment-related anxiety, self-esteem, and social anxiety were intercorrelated. A mediation analysis conducted using the *PROCESS* v4.0 macro for SPSS, found that the indirect effect of attachment-related anxiety on social anxiety through self-esteem was significant. This finding is congruent with a theoretical account linking attachment-related anxiety to social anxiety through the mediating role of the internal working model of the self.

## Introduction

Social anxiety is characterised by a fear of negative evaluation by others.(1) Most people experience some degree of social anxiety in certain situations, such as job interviews or first dates.(2) However, for some people, social anxiety occurs more frequently and may impact even mundane decisions, to the point of avoiding certain social situations or experience intense discomfort during them.(3) A higher degree of social anxiety is associated with less satisfying experiences in romantic (4) and peer (5) relationships, as well as being viewed as less likeable by others.(6) It is also a risk factor for depression.(7)

Social anxiety symptoms exist on a continuum, and people who experience social anxiety may, or may not, meet the clinical criteria for Social Anxiety Disorder (SAD).(8) In Europe, the median 12-month prevalence of SAD is 2%,(9) and it disproportionately affects young people and females.(10) SAD usually emerges during childhood or adolescence.(11)

Whilst social anxiety has been widely researched, its aetiology is not yet fully understood.(12) One widely researched framework for improving our understanding of social anxiety is that of attachment theory. As indicated in a recent systematic review, adult attachment is associated with social anxiety.(13) Experiences in attachment relationships may contribute to social anxiety by shaping people’s views of themselves and others.(14) This study tested an aspect of this claim by examining whether self-esteem mediated the relationship between attachment-related anxiety and social anxiety in adults. If this is the case, then interventions aimed at improving self-esteem may decouple attachment-related anxiety and social anxiety, and, therefore, be a viable method of treating social anxiety.

### Adult Attachment and Social Anxiety

Attachment theory could improve our understanding of the aetiology of social anxiety, through its theory of internal working models (IWMs) of the self and others. IWMs – a key explanatory tool within attachment theory – may explain the development of the views of the self and others that underly social anxiety.(14)

Attachment theory emerged in the second half of the 20^th^ century with the publication of seminal works by Bowlby (15–17) and Ainsworth.(18) Bowlby argued that infants are innately endowed with a system to facilitate the forming of close bonds with their primary caregiver(s).(19) This system gives rise to instinctive behaviours, such as proximity-seeking, that promote the development of attachment bonds.

Interactions between the infant and the caregiver can be shaped by the degree of the caregiver’s sensitive responsiveness to these behaviours.(18) Repeated patterns of interaction in infant-caregiver dyads are believed to be gradually internalised by the infant, leading to different possible patterns of attachment-related behaviour in future interactions.(17) Ainsworth and colleagues proposed that these patterns could be sorted into secure and insecure (avoidant and anxious-ambivalent) categories.(18) Securely attached infants are believed to use their caregiver as a secure base from which to venture into the world, and to return to in the face of stressful situations. Avoidant infants experience anxiety about the caregiver’s responsiveness and therefore display detachment from the caregiver in unfamiliar contexts. Anxious-ambivalent infants appear distressed in unfamiliar contexts and display attachment-seeking behaviours but may appear angry and inconsolable when reunited with the caregiver.(18) Subsequent longitudinal research has demonstrated that infant attachment security predicts future mental health and interpersonal functioning.(20–22)

Researchers have conceptualised adult romantic love using the framework of attachment theory.(23) For example, Hazan and Shaver (23) argued that adult romantic love is an attachment process, and that patterns of attachment in romantic relationships mirror Ainsworth’s typology of attachment styles in infant-caregiver dyads.(18,23) However, subsequent research has shown that adult attachment patterns can be understood as dimensional, not categorical.(24,25) In addition, factor analytic studies have demonstrated that self-reported adult attachment may reflect both attachment-related anxiety and attachment-related avoidance.(26) These two factors may play a role in explaining the development of social anxiety.

These anxiety and avoidance dimensions have been conceptualised as two subsystems of the attachment system.(27,28) The attachment-related anxiety subsystem monitors and evaluates the likelihood of achieving attachment-related goals, such as the reciprocation of feelings for a romantic partner. The attachment-related avoidance subsystem regulates attachment-promoting behaviours, for example seeking proximity to attachment figures.

Individual differences in attachment-related anxiety and avoidance scores reflect variation in the functioning of these two subsystems.(27) An above-average score on attachment-related anxiety indicates a high degree of concern about one’s ability to meet attachment-related goals whereas a high score on attachment-related avoidance reflects a tendency to avoid proximity to attachment figures.(27)

A number of studies have found positive associations between social anxiety and attachment-related anxiety and avoidance. In a systematic review, Manning and colleagues(13) examined 30 studies that tested for correlations between adult attachment and social anxiety. 28 of the studies found that insecure attachment and social anxiety were positively correlated.(13) Subsequent research has also found this to be the case.(29,30) Some earlier studies that investigated the relationship between adult attachment and social anxiety used categorical measures of attachment,(31,32) which fail to reflect the latent structure of individual differences in adult attachment.(24,27) However, most recent studies have used variants of the Experiences in Close Relationships Scale (ECR),(26) which is a valid measure of the anxiety and avoidance dimensions.(27,33) That said, only one study has specifically investigated the role that the two dimensions play in predicting social anxiety symptoms.(30) This study aimed to extend this limited research by exploring the relationship between attachment-related anxiety and social anxiety.

### The Internal Working Model of the Self

Whilst previous studies have reported associations between adult attachment and social anxiety, little attention has been paid to explaining this relationship. The current study contends that attachment-related anxiety and avoidance may contribute to the development of social anxiety through the mechanism of IWMs of the self and others.(14) IWMs are mental representations of interactional patterns with attachment figures which guide interpretations of, and predictions about, interpersonal events, as well as decisions as to how one should interact.(34) These models are hypothesised to evolve gradually during infancy as a result of repeated patterns of interaction occurring within infant-caregiver dyads.(15) They are conceived as dynamic, meaning that they are the basis for mental simulations of interactional patterns.(34) Although Bowlby argued that they function automatically and outside of conscious awareness,(17) elsewhere he acknowledged that they include an affective element, which allows for the appraisal of the self and others as good or bad.(15,34)

Bowlby held IWMs of the self and others to be the mechanism by which infant attachment styles affect later personality, relationship patterns, and interpersonal functioning.(15) Given that social anxiety occurs within interpersonal contexts and is a function of one’s expectations about oneself and others,(35) it is plausible that negative IWMs of the self and/or of others shape these expectations and thus contribute to the development of social anxiety.(13,14)

If IWMs contribute to social anxiety in the manner just outlined, then they may explain the relationship between attachment-related anxiety and social anxiety. Sustained anxiety about an attachment figure’s continued love or availability (attachment-related anxiety) may lead to a view of the self as unworthy of attention and support, i.e., a negative IWM of the self.(14,36) This negative IWM of the self may, in turn, contribute to the development of social anxiety.(35) By virtue of this reasoning, this study argues that the IWM of the self may explain the relationship between attachment-related anxiety and social anxiety.

### The Mediating Role of Self-Esteem

In this study, it was hypothesised that self-esteem mediated the relationship between attachment related anxiety and social anxiety. Self-esteem is understood as the individual’s overall judgement of themselves as good or bad.(37) It is believed to incorporate both cognitive and affective elements.(38,39) Additionally, it has been argued that self-esteem functions within a relational context as a ‘sociometer’: a gauge of the degree of a person’s perceived inclusion or exclusion by others.(40) Thus, both self-esteem and the IWM of the self are argued to indicate the individual’s evaluation of their own self-worth in a relational context, suggesting an overlap between the two constructs. With this in mind, the IWM of the self was operationalised by measuring participants’ self-esteem.

The current study was not the first to operationalise the IWM of the self in this way. Applying similar theoretical grounding, Seon(41) tested the relationship between insecure attachment, self-esteem and social anxiety in a sample of Korean children. As in the present study, self-esteem was held as an appropriate measure of participants’ IWMs of the self.

Seon’s(41) findings indicated that self-esteem mediated the relationship between the other two variables. However, the current study was the first to test this hypothesis in an adult sample.

Research indicates close ties between attachment-related anxiety, social anxiety, and self-esteem in adults. Leary and colleagues’(35,42) self-presentation theory posits that social anxiety occurs when an individual is motivated to make a particular impression on an audience that is perceived as important, but doubts that they will succeed in doing so. Self-esteem is viewed as a gauge of the individual’s perceived inclusion or exclusion by important others, with low self-esteem indicating that the individual considers themselves likely to be excluded(35,40). This belief may in turn produce an increase in the individual’s level of social anxiety(35). This account is empirically supported. For example, self-esteem is negatively associated with both SAD and social anxiety symptoms.(43–45) Additionally, Baldwin and Main(46) found that cueing participants to think about being socially rejected led to increased social anxiety, suggesting that social anxiety may be the output of an internal sociometer.(35)

Self-esteem is also negatively associated with attachment-related anxiety.(47–49) Furthermore, a meta-analysis of longitudinal studies found that self-esteem and social relationships reciprocally influence each other.(50) This is congruent with the idea that attachment-related experiences affect self-esteem. Together, the findings discussed constitute grounds for the hypothesis that self-esteem mediates the relationship between attachment-related anxiety and social anxiety.

### Purpose of the Present Study

This study sought to explore one possible explanation for the relationship between attachment-related anxiety and social anxiety, namely the mediating role of the IWM of the self. The IWM of the self was operationalised by measuring participants’ self-esteem. The research questions were as follows: 1) are attachment-related anxiety and social anxiety associated? and 2) does self-esteem mediate the relationship between attachment-related anxiety and social anxiety? The implications of attachment theory and the explanatory importance of IWMs within it led to the following hypotheses: 1) attachment-related anxiety is positively associated with social anxiety, 2) attachment-related anxiety is negatively associated with self-esteem, 3) social anxiety is negatively associated with self-esteem, and 4) self-esteem mediates the relationship between attachment-related anxiety and social anxiety. These hypotheses were tested by means of an anonymous, quantitative survey. The study was intended to improve our understanding of the relationship between adult attachment and social anxiety, specifically by exploring whether the IWM of the self may explain this relationship. If this is the case, then interventions aimed at improving self-esteem may decouple attachment-related anxiety and social anxiety, and therefore be a viable method of treating social anxiety.

## Methods

### Participants

Participants were adults over the age of 18 sampled from the general population (*N* = 63, 79.4% female). All participants were residents of the United Kingdom at the time of participation. Table 1 shows the sociodemographic characteristics of the participants.

**Table 1.** Sociodemographic Characteristics of Participants.

| Factor | n | % |
| --- | --- | --- |
| Gender |  |  |
| Male | 13 | 20.6 |
| Female | 50 | 79.4 |
| Age group |  |  |
| 18-24 | 44 | 69.8 |
| 25-34 | 12 | 19 |
| 35-44 | 3 | 4.8 |
| 45-54 | 1 | 1.6 |
| 55-64 | 3 | 4.8 |
| Relationship status |  |  |
| Married | 4 | 6.3 |
| In a relationship | 21 | 33.3 |
| Divorced | 1 | 1.6 |
| Single | 33 | 52.4 |
| Never been in a relationship | 3 | 4.8 |
| Prefer not to say | 1 | 1.6 |
| Ethnic background |  |  |
| White/Caucasian | 23 | 36.5 |
| Asian/Asian British | 38 | 60.3 |
| Other | 1 | 1.6 |
| Prefer not to say |  | 1.6 |

## Measures

### Demographic Questionnaire

Participants were asked to report their gender, age group, relationship status and ethnic background.

### Anxiety Subscale of the Experiences in Close Relationships Revised Scale (ECR-R)(***51***), Shortened

To measure their degree of attachment-related anxiety, participants completed a shortened version of the Anxiety subscale of the ECR-R. The ECR-R was designed to capture the two dimensions of adult attachment – anxiety and avoidance(51) – whereas some alternative instruments incorrectly assume that adult attachment can be categorised into discrete styles.(23) The measure is more predictive of variance in anxiety and avoidance in adult romantic relationships (the focus of this study) than within family relationships or friendships.(52) It has high test-retest reliability(33) and internal consistency(51) and is widely used in attachment research.(53)

Participants responded to statements on a seven-point Likert scale, ranging from 1 = *Strongly Disagree* to 7 = *Strongly Agree*. The statements were intended to test participants’ degree of attachment-related anxiety within their romantic relationships, with a higher score indicating a higher degree of attachment-related anxiety. The statements included: “I’m afraid that I will lose my partner’s love” and “I worry a lot about my relationships”. A factor analysis conducted by Sibley and colleagues was used to shorten the subscale, in order to reduce the amount of time needed to complete the survey.(52) The full subscale has 18 items, and it was shortened by removing the nine items that loaded least strongly on the Anxiety subscale, leaving nine items in total. In this study, the remaining nine items, which were the instrument used to measure attachment-related anxiety, had an internal consistency of .94 (Cronbach’s alpha).

### Social Interaction Anxiety Scale (SIAS).(***54***)

The SIAS was used to assess participants’ general social anxiety, or fear of and during social interaction. The SIAS is commonly used in social anxiety research as a measure of the fear of interaction in generic social situations, and the items reflect anxiety in a variety of different social interactional contexts.(55,56) The SIAS has high test-retest reliability and internal consistency, as well as good convergent validity.(54)

The SIAS is a 20-item Likert scale which requires participants to endorse statements about their cognitive, emotional, and behavioural responses to different social scenarios, such as meeting new people. Responses are scored from 0 to 4, where 0 = *Not at all* and 4 = *Extremely*. Three of the items have to be reverse-scored, and a higher score indicates a higher degree of social anxiety. The statements include: “I have difficulty making eye contact with others” and “I am nervous mixing with people I don’t know well”. Cronbach’s alpha in this study was .90.

### Rosenberg Self-Esteem Scale (RSES).(***37***)

The RSES was used to measure participants’ degree of self-esteem. The scale has good test-retest reliability(57) and internal consistency.(58) Furthermore, Trzesniewski and colleagues found that RSES scores in adolescents predicted subsequent mental health outcomes, indicating that the instrument is predictively valid, as well as relevant to this study.(59)

The RSES is a four-point, ten-item Likert scale in which statements are rated from *Strongly Agree* to *Strongly Disagree*. Five of the ten items have to be reverse-scored, and a higher score indicates a higher degree of self-esteem. The statements include: “On the whole, I am satisfied with myself” and “I feel I do not have much to be proud of”. In this dissertation, Cronbach’s alpha was .85.

## Procedure

Participants were recruited via the University of Edinburgh’s online learning portal and social media platforms, as well as through word-of-mouth. A flyer with a link to the study was shared on these platforms along with a standard message explaining the purpose of the study, the inclusion criteria, and the contact details of one of the researchers. The participants followed a link to the questionnaire, which was created using the Online Surveys platform. They were required to read a Participant Information Sheet and asked to consent to participate in the study, and to their data being utilised by the researchers. They were also debriefed after completing the survey and directed to mental health resources. No personally-identifiable information was collected, and a numerical ID was allocated automatically to each participant, ensuring anonymity and confidentiality. Ethical approval to conduct this study was given by the University of Edinburgh School of Health in Social Science Research Ethics Committee (5^th^ May 2022).

### Quantitative Analyses

All analyses were carried out using IBM SPSS Statistics 25. Hypotheses 1-3 (that attachment-related anxiety, self-esteem and social anxiety would be intercorrelated) were tested with bivariate Pearson correlations. Multiple regression analyses were used to test whether the relationships between each of the pairs of variables remained significant when controlling for the third variable. Significance was tested at the α = .05 and α = .01 levels for the Pearson correlations and the multiple regression analyses. Following Armstrong’s(60) recommendations, the Bonferroni correction was not applied to adjust for multiple tests. The mediation analysis required to test hypothesis 4 was conducted using the *PROCESS* v4.0 macro.(61) The indirect effect was estimated by entering social anxiety as the outcome variable, attachment-related anxiety as the independent variable, and self-esteem as the mediating variable, then selecting Model 4.

The bootstrap method for testing mediation was used. The use of bootstrapping as a method of estimating the precision of the estimates is advantageous because it does not assume that the sampling distribution of the parameter being estimated is normally distributed.(62) This is valuable in this context because clinical constructs, such as social anxiety, often have skewed distributions in normal populations,(62) and because the small sample size may have led to reduced power to detect a deviation from normality.(63) The bias-corrected bootstrap method has been shown to have higher power than other approaches, including Baron and Kenny’s traditional method for testing mediation,(64) which is advantageous given the small sample used in this study.(65)

The 95% confidence intervals of the indirect effect were estimated using the bias-corrected accelerated bootstrap, with 5000 bootstrap samples requested. This method produces a range of values within which the effect size of the indirect effect would lie in 95% of studies. If the 95% confidence intervals do not contain zero, then this indicates that in 95% of studies, the indirect effect would not be zero. This finding supports the rejection of the null hypothesis. Conversely, if the confidence intervals contain zero, then the null hypothesis cannot be rejected.(63)

### Power Analysis

To estimate the required sample size, an a priori power analysis was conducted using Schoemann and colleagues’ online application: Monte Carlo Power Analysis for Indirect Effects.(66) This method was selected because it is preferable to analytical methods, such as those implemented in G*Power, when estimating the indirect effect of a mediation model using bootstrapping.(67) Target power was not achieved, however, due to a slow response rate and limited time.

## Results

### Descriptive Statistics

Total scores on the three scales were computed by summing the participants’ scores on each of the items. Due to the small sample size, missing cases were replaced with the series mean (i.e., the mean of all the values in the variable) in order to retain power. No outliers were identified. Table 2 shows the means, standard deviations, minimums, and maximums of each scale.

**Table 2.** Correlations, Means, Standard Deviations, Minimums, and Maximums.

|  | Correlations |  | Mean | SD | Min | Max |
| --- | --- | --- | --- | --- | --- | --- |
|  | 2 | 3 |  |  |  |  |
| Attachment anxiety | -.44** | .41** | 30.59 | 14.59 | 10 | 63 |
| Self-esteem |  | -.53** | 17.65 | 5.08 | 5 | 27 |
| Social anxiety |  |  | 33.79 | 13.98 | 8 | 68 |

\*\* $p < .01$

### Analyses

The assumptions of the linear model were met.(63) That is, the outcome variable (social anxiety) was linearly related to both predictors (attachment-related anxiety and self-esteem). A Durbin-Watson test indicated that the errors were uncorrelated. To test for homoscedasticity, the standardised predicted values were plotted against the standardized residuals; no pattern was visible, indicating that the variance of the standardised residuals was constant. A normality test indicated that the standardised residuals were normally distributed. Finally, the Pearson correlation tests indicated that multicollinearity fell below the permissible threshold.(68)

#### Hypotheses 1-3

Pearson correlation tests were used to assess the associations between the three variables; table 2 shows the zero-order correlations between the variables. The relationship between attachment-related anxiety and social anxiety was assessed. As hypothesised, the Pearson correlation test showed that the two were positively correlated, *r* = .41, *p* = .001. The relationship between attachment-related anxiety and self-esteem was also assessed. As hypothesised, the Pearson correlation test showed that the two were negatively correlated, *r* = -.44, *p* = .000. Finally, the relationship between social anxiety and self-esteem was assessed. As hypothesised, the Pearson correlation test showed that the two were negatively correlated, *r* = -.53, *p* = .000. All of these correlations were significant at the α = .01 level.

Given the demographics of the sample (see Table 1) post-hoc exploratory tests were undertaken to check ethnicity and gender as potential moderators, but no significant effects were found. Post-hoc multiple regression analyses with enter method were also conducted to test whether each of these relationships remained significant when controlling for the third variable. Table 3 shows the three multiple regression models. Attachment-related anxiety and social anxiety no longer predicted each other when controlling for self-esteem, which is congruent with hypothesis 4, that self-esteem mediates the relationship between the other two variables.

**Table 3.** Results of Multiple Regression Analyses Predicting Social Anxiety, Attachment-Related Anxiety and Self-Esteem.

| Dependent variable | Predictors | *B* | *SE b* | $\beta$ | *p* |
| --- | --- | --- | --- | --- | --- |
| Social anxiety | Attachment anxiety | .21 | .11 | .22 | .066 |
|  | Self-esteem | -1.18 | .33 | -.43 | .001** |
| Attachment anxiety | Social anxiety | .26 | .14 | .25 | .066 |
|  | Self-esteem | -.89 | .38 | -.31 | .023* |
| Self-esteem | Attachment anxiety | -.09 | .04 | -.27 | .023* |
|  | Social anxiety | -.15 | .04 | -.42 | .001** |

\*\* $p < .01$ \* $p < .05$

#### Hypothesis 4

Figure 1 shows the mediation model and its unstandardised beta coefficients. Table 4 shows the unstandardised and standardised beta coefficients of the mediation model. The total effect model explained a statistically significant amount of the variance in social anxiety, *F*(1, 61) = 12.45, *p* = .001, *R^2^* = .17. The total effect (*c*) of attachment-related anxiety on social anxiety was significant, *b* = .43, *β* = .41, *t*(60) = 3.53, *p* = .001. The direct effect (*c’*) of attachment-related anxiety on social anxiety was not significant, *b* = .26, *β* = .25 *t*(60) = 1.87, *p* = .066. The indirect effect (*ab*) of attachment-related anxiety on social anxiety through self-esteem was significant, *b* = .17, *β* = .16, 95% BCa CI [.01, .37]. The indirect effect accounted for 39% of the association between attachment-related anxiety and social anxiety (*β* _indirect effect_ / *β* _total effect_, i.e., .17/.43). The significant indirect effect indicates that, as hypothesised, self-esteem mediates the relationship between attachment-related anxiety and social anxiety in this sample.

**Fig 1.**
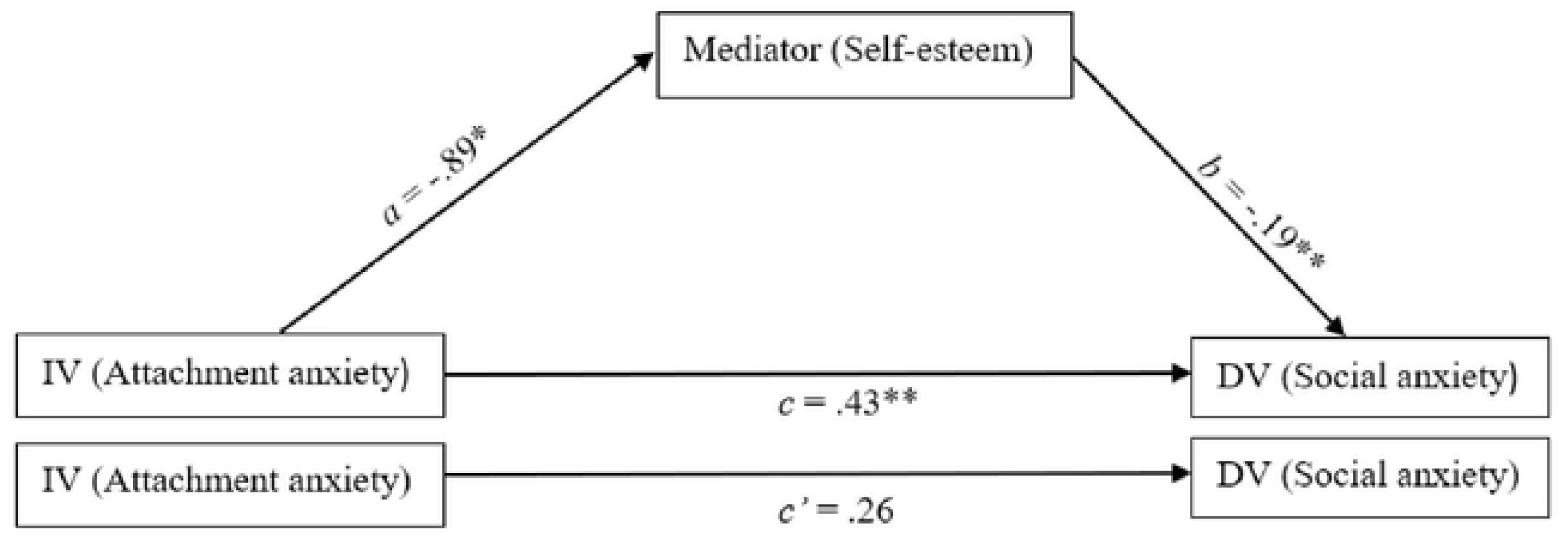
Unstandardised beta coefficients for the mediation model with social anxiety as the dependent variable ** p < .01, * p < .05

**Table 4.**
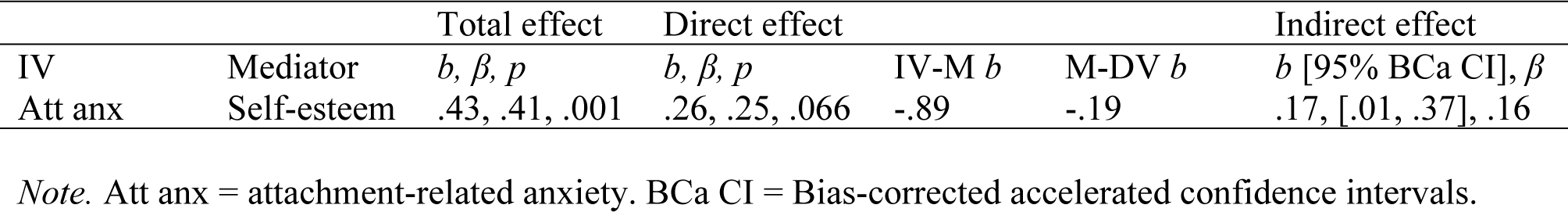
The Beta Coefficients of the Mediation Model with Social Anxiety as the Dependent Variable.

## Discussion

This study tested a mediation model with self-esteem as mediator in the relationship between attachment-related anxiety and social anxiety. All three of the variables were intercorrelated in the hypothesised directions. Multiple regression analyses indicated that attachment-related anxiety and social anxiety did not predict each other when controlling for self-esteem, but that self-esteem predicted both attachment-related anxiety and social anxiety. However, this was a post-hoc exploratory analysis and should therefore be treated with caution.(69)The 95% confidence intervals of the indirect effect of attachment-related anxiety on social anxiety did not contain zero, indicating that the null hypothesis could not be kept.(70) Thus, in all four cases, the results supported the rejection of the null hypotheses. It should be noted that females, Asians and young people (18–24) were overrepresented in the study sample. These findings may therefore not be generalisable to the rest of the population.

The fact that attachment-related anxiety, social anxiety, and self-esteem were all intercorrelated coheres with previous findings.(13,43–45,48,49). This study built on these findings by demonstrating that self-esteem mediates the relationship between attachment-related anxiety and social anxiety. Whilst self-esteem has been found to mediate insecure parental attachment and social anxiety in Korean children,(41) this study extended this finding to romantic attachment in an adult British population. The results hereby presented are congruent with a theoretical account linking attachment-related anxiety to social anxiety through the mechanism of the IWM of the self.(14) Individuals who score high on attachment-related anxiety have a high degree of anxiety about their ability to meet attachment-related goals.(27) This may lead to the construction of a model of the self as unworthy of attention and support.(71) Viewing oneself in this way may, in turn, lead to increased social anxiety(14). However, due to the observational and cross-sectional design of the study, causality was not established.

One of Kazdin’s (72) criteria for inferring the occurrence of a causal process is that this process is predicted by a broader scientific theory. A strength of this study is therefore that the mediation hypothesis was derived from a theoretical account of the effects of attachment experiences on later outcomes, which can be, and has been, applied to the development of social anxiety.(14) The IWM construct is a well-established and widely researched element of attachment theory, and there is agreement amongst attachment researchers as to its explanatory importance.(34,73) Moreover, Vertue’s (14) unifying theory of the development of social anxiety held IWMs to be the crucial mechanism underlying it. It is therefore theoretically plausible that the IWM of the self is the mechanism that links attachment-related anxiety and social anxiety.

The findings of this study raise questions that could be explored in future research. First, one should consider how self-esteem relates to other mediators of the relationship between adult attachment and social anxiety that have been identified. These include cognitive variables such as emotion regulation strategies,(30) cognitive flexibility,(74) locus of control, and repetitive thinking (75) evolutionary variables, namely social comparison and submissive behaviour (76); depression (77); hope (78); social approach and avoidance motivation (79); and perceived social support.(80) Manning and colleagues (13) note that the overlap or association between these variables is unknown. It is therefore plausible that some of these significant indirect effects are the result of the variables being confounded with one another. For example, depression is negatively associated with self-esteem.(81) Future process research with relation to attachment and social anxiety could explore this possibility.

A second question raised by this study is whether its logic could be applied to attachment-related *avoidance* and its relationship to social anxiety. Manning and colleagues’ (13) review indicates that the two constructs are about as strongly associated as attachment-related anxiety and social anxiety. Whilst attachment-related anxiety has been linked to a negative IWM of the self, it is argued that attachment-related avoidance relates to negative IWMs of others.(71,82) It could therefore be argued that attachment-related avoidance leads to social anxiety through the mechanism of negative IWMs of others.

### Limitations

This study had several limitations. First, the sample was relatively small, and the primary disadvantage of a small sample size is that it increases the likelihood of a Type II error, that is, the failure to reject the null hypothesis when it is false.(83) The target power level of .80 was not achieved, meaning that the likelihood of a Type II error was higher than 20%.(84) Despite this, the null hypotheses were indeed rejected, and our findings are congruent with the effects hypothesised in the introduction.

Second, the instrument used to measure attachment-related anxiety was a shortened non-validated version of one subscale from the ECR-R, in favour of a shorter procedure. Internal consistency was high (α **=** .94), indicating that the instrument measured a single construct; however, it is possible that removing some items led to reduced reliability or validity. This issue could be addressed in future research by using already validated measures of attachment-related anxiety, such as the full ECR-R, or the Experiences in Close Relationships Scale – Short Form.(85)

Third, the data collected for this study were of a cross-sectional nature. An essential criterion for establishing that a mediator plays a causal role is that the predictor, outcome, and mediator variables exist in appropriate temporal relation to each other.(72) Specifically, the mediator should precede the dependent variable, and the independent variable should precede both the mediator and the dependent variable. This cannot be established using cross-sectional data, and this study therefore does not show that attachment-related anxiety leads to social anxiety through the mechanism of self-esteem. This is a critique that can be levelled at most of the past studies examining potential mediators of the relationship between adult attachment and social anxiety.(30,74–80)

To examine the temporal relationships between adult attachment, social anxiety, and potential mediators, future research should use longitudinal methods. One study has successfully examined the longitudinal relationship between adult attachment and later social anxiety.(31) In this study, participants responded to questionnaires at two time points, and insecure adult attachment at Time 1 was found to predict social anxiety at Time 2. However, given the dimensional nature of adult attachment,(25,51) the study’s use of a categorical instrument to measure attachment limits the value of this finding.(13) More longitudinal research is therefore warranted.

A final limitation of this study is that all of the variables were measured using self-report instruments. Utilising instruments of this nature introduces the risk of bias due to various method effects.(86) However, this risk is less significant in cases when the construct validity of the variable(s) in question has been established, for example by testing for convergent and discriminant validity.(86) Research has supported the convergent and discriminant validity of the instruments utilised in this study.(52,56,87) Therefore, this risk of bias should not be overemphasised. A more important aspect of this limitation relates specifically to the use of self-reported self-esteem as a measure of the IWM of the self. IWMs are posited to operate outside of conscious awareness,(17) meaning that any self-report instrument would be unlikely to accurately capture these processes.(73) It is plausible that the self-esteem measure tracks the affective element of the IWM of the self, i.e., as either good or bad,(34) however this is only one aspect of the overall construct.

### Future directions

This study did not control for potentially confounding variables, apart from ethnic background and gender. For example, Aderka and colleagues found that, when controlling for submissive behaviour and social comparison, adult attachment no longer predicted social anxiety.(76) That said, although Aderka and colleagues used the ECR to measure attachment, they did not distinguish between the attachment-related anxiety and avoidance factors.(76) Other potential confounds include depression(81,88) and personality variables such as neuroticism.(89–91) Future research should control for these variables to test whether the relationship between social anxiety and attachment-related anxiety is robust.

Future research could also use implicit methods as a means of accessing the unconscious processes at the root of IWMs.(73) For example, in one study, participants’ working models were inferred from their written interpretations of relationship events.(92)

### Implications for Clinical Practice

Current best practices for treating SAD include cognitive-behavioural therapy (CBT) and pharmacotherapy.(93) Conceptual research, such as reported in this paper, can inform the application of CBT to case-conceptualisations for social anxiety. Clinicians may wish to explore the role that experiences in attachment relationships, and attachment-related anxiety, play in leading to or maintaining social anxiety symptoms. They may also consider the IWM of the self and/or self-esteem as factors underlying the development of social anxiety. A concrete implication is that intervening to improve self-esteem may be a viable method of reducing social anxiety, although further research would be required to establish a causal link between self-esteem and social anxiety. For example, in line with Rapee and Heimberg’s model, cognitive-behavioural therapists may attempt to guide patients with SAD in restructuring negative cognitive representations of the self.(94) In this way, social anxiety symptoms could be decoupled from the attachment-related anxiety that may underlie them.

### Conclusion

This study confirmed significant associations between attachment-related anxiety, social anxiety, and self-esteem in a predominantly young, female and Asian sample. This study built on this finding by testing whether self-esteem mediated the relationship between attachment-related anxiety and social anxiety; our statistical analyses indicate that mediation did occur. Although the data were cross-sectional, the findings are congruent with a theoretical account linking attachment-related anxiety to social anxiety through the mechanism of a negative IWM of the self. Further research and longitudinal study designs are warranted to explore the role that IWMs may play in linking attachment and social anxiety, and adding an indirect measurement of IWMs is recommended.

## Data Availability

The data will be stored by the respository DataShare at the University of Edinburgh. URLs/accession numbers/DOIs will be available after the acceptance of the manuscript for publication.

## Acknowledgements

We would like to express gratitude to the participants in this study, for the time they put into completing our survey.

We would also like to thank Ye Tang, Ke Ju, Jiawei Wang, and Jessie Kan, who contributed to the process of creating our survey, and writing our research proposal and ethics application, as well as contributing their thoughts and ideas about this study.

